# Care-seeking pathways and time to presentation for stroke care at UNIMEDTH, Ondo State, Nigeria

**DOI:** 10.64898/2026.06.04.26354906

**Authors:** Olubusayo Ogunsemoyin, Olufunke Fayehun

## Abstract

**Introduction:** Stroke care is time-sensitive, yet patients in low-resource settings may reach tertiary services only after passing through multiple formal and informal care options. This study examined documented care-seeking pathways and time to presentation among stroke cases recorded at the University of Medical Sciences Teaching Hospital (UNIMEDTH), Ondo State, Nigeria.

**Methods:** A retrospective hospital record review was conducted using secondary data from the Stroke Registry, radiology department records, referral notes, and ambulance records at UNIMEDTH. The analysis included 371 stroke cases with documented time from symptom onset to UNIMEDTH presentation and reconstructable care pathways. First-contact routes were classified as hospital/biomedical, self/informal or traditional/faith-based care, and the number of documented steps defined pathway complexity before and including tertiary presentation. Frequencies and percentages described pathway patterns; median presentation times were compared using Mann-Whitney U and Kruskal-Wallis tests.

**Results:** The median time to tertiary presentation was 24 hours (interquartile range [IQR] 9-72), and 317 patients (85.4%) presented after four hours. Only 30 patients (8.1%) presented directly to UNIMEDTH; 44 distinct care-pathway sequences were recorded. Hospital-facility first contact was documented for 81 patients (21.8%). It was associated with a median presentation time of 3 hours (IQR 2-6), compared with 48 hours (IQR 24-72) among patients whose initial contact was outside a hospital facility (U = 699.50, p < 0.001). The median time also differed across grouped first-contact categories and pathway complexity levels (both p < 0.001).

**Conclusion:** Non-hospital or multi-step care-seeking pathways commonly preceded tertiary stroke presentations in this setting. The findings indicate that delayed tertiary arrival is partly embedded in the pathway followed after symptom onset. Interventions should combine public recognition of stroke warning signs with urgent referral linkages involving hospitals, patent medicine vendors, traditional and faith-based providers, and emergency transport systems.

## Introduction

Stroke remains a major cause of death, disability and long-term dependency worldwide [1-3]. Global Burden of Disease 2021 estimates indicate 11.9 million incident strokes and 7.3 million stroke deaths in 2021, with substantial disparities across regions and levels of socioeconomic development [1-3]. In low- and middle-income countries (LMICs), the burden is exacerbated by uneven access to diagnostic imaging, organised acute stroke services, emergency transport and post-stroke rehabilitation [4-7].

Intravenous alteplase is a cornerstone treatment for eligible patients with acute ischaemic stroke. It has been shown to improve functional outcome when given within 4.5 hours of symptom onset, with earlier treatment yielding greater benefit [8-10]. Later reperfusion options depend on specialised assessment and imaging capacity that may not be routinely available in many low-resource settings [11]. Thus, early arrival at a capable hospital remains an essential precondition for time-dependent treatment, even when eligibility cannot be determined solely from arrival time.

Evidence from across Africa indicates that late hospital presentations are common. A systematic review of stroke care in Africa reported a median onset-to-admission time of 31 hours and identified limited symptom recognition, transport constraints, and inadequate organised stroke services as persistent barriers [7]. Single-country data show similarly long delays: median onset-to-door of 20 h in Mozambique [15], 24 h in South Africa, and very high late-arrival prevalence (≈90%) in Uganda, with about 50% in Sierra Leone [16,17]. In Sierra Leone, the first use of other providers before referral to the hospital added about 38 extra hours [17]. African review data link delays in data collection to late referral from private hospitals, visits to traditional healers, and treatment at home [8]. A subsequent meta-analysis likewise found a high burden of prehospital delay among stroke patients in Africa, with poor knowledge of warning signs and greater distance from health facilities contributing to delayed arrival [15-19]. The World Stroke Organisation has therefore emphasised that strengthening prehospital stroke care in LMICs requires attention to community recognition, emergency transport, referral coordination, and first-contact providers outside specialised hospitals [20].

African evidence shows that first-contact choice and pathway complexity (multiple sequential providers) are major drivers of onset-to-tertiary delay, beyond initial help-seeking speed [1, 14-16]. In pluralistic systems like Nigeria, similar patterns are likely: movement between informal providers, clinics, and hospitals before reaching a stroke-capable tertiary centre is expected to inflate referral delays.

Existing stroke-delay studies have largely focused on the prevalence and correlates of late arrival. While important, these approaches may not fully reveal the sequence in which time is lost. A patient may seek help promptly after symptom onset yet still reach tertiary stroke care late if the initial contact does not initiate rapid referral. This study, therefore, examines documented care-seeking pathways among stroke patients presenting at UNIMEDTH in Ondo State, Nigeria, and assesses how time to tertiary hospital presentation varies by first point of contact and pathway complexity.

### Theoretical framework

The study is organised primarily around a pathways-to-care perspective, which treats care seeking as an ordered process from symptom onset through initial contact and subsequent transitions to a definitive facility. Rather than reducing help seeking to a single choice, this perspective directs attention to the providers encountered, the sequence of transitions, and the time that may accumulate between them. Such an approach is especially relevant for acute stroke, because contact with any provider is not equivalent to timely entry into a service capable of urgent stroke assessment. MacKian argues that attention to care routes and provider transitions improves the policy value of health-seeking research, particularly in settings where people move between multiple systems of care [21, 22].

Two complementary perspectives support the interpretation of the observed pathways. Andersen’s Behavioural Model of Health Service Use conceptualises service utilisation as shaped by predisposing characteristics, enabling resources and need [23-25]. In the present analysis, it helps frame first contact as potentially influenced by provider accessibility, household resources, transport, and perceived severity. However, the study does not estimate the independent effects of these determinants. Kleinman’s explanatory model further recognises that symptoms are interpreted within cultural and social worlds, and that therapeutic choices may therefore include biomedical, traditional or faith-based responses [26]. The present study uses these perspectives to interpret pathway patterns without treating any recorded route as evidence of irrationality or as proof of causal delay.

Operationally, the framework is represented by two dimensions: first-contact category and pathway complexity. First contact distinguishes hospital/biomedical care, self/informal care and traditional/faith-based care, while pathway complexity records the number of documented contacts culminating in UNIMEDTH presentation. Together, these measures allow examination of whether the route taken before tertiary arrival is associated with the observed time to presentation.

## Materials and methods

### Study design and setting

This study was a retrospective hospital-based review of stroke cases presenting at the University of Medical Sciences Teaching Hospital (UNIMEDTH), Ondo State, Nigeria. UNIMEDTH is a tertiary facility that receives patients through direct presentation and referrals from other providers and facilities. The study was reported in accordance with the STROBE guidance for observational studies [27].

### Data sources and study population

Secondary data were obtained from the UNIMEDTH Stroke Registry, radiology department records, referral notes, and ambulance records. These sources were used to identify recorded stroke cases and reconstruct documented care-seeking pathways for the 24 months preceding the study. The data were accessed for research purposes from 22 July 2024 to 17 January 2025.

The analytical sample comprised 371 unique stroke cases with available sociodemographic information, recorded time from symptom onset to arrival at UNIMEDTH, and a reconstructable care pathway. The analysis, therefore, includes cases that reached UNIMEDTH and for whom relevant pathway information was documented.

### Measurement of variables

The primary outcome was time-to-presentation for stroke care at the tertiary hospital, measured in hours from recorded symptom onset to arrival at UNIMEDTH. Presentation time was analysed as a continuous, right-skewed duration and summarised using medians and interquartile ranges (IQRs). A descriptive binary timing category classified presentations as early (occurring within 4 hours) and late (occurring after 4 hours). This operational cut-off identifies an early-arrival group in the record dataset; it should not be interpreted as establishing clinical eligibility for thrombolysis or any other acute intervention.

Care pathway variables were derived from documented contacts between symptom onset and arrival at UNIMEDTH. The first points of contact included UNIMEDTH, government hospital, private hospital, primary health centre, management at home by a doctor or nurse, self-medication, chemist or patent medicine vendor, traditional home remedy, traditional healer and prayer house. For the grouped classification, hospital/biomedical care included UNIMEDTH, government hospital, private hospital, primary health centre and management at home by a doctor or nurse; self/informal care included self-medication and use of a chemist or patent medicine vendor; and traditional/faith-based care included traditional home remedy, traditional healer and prayer house.

A second first-contact measure distinguished hospital-facility contact from non-hospital contact. In this comparison, contacts with UNIMEDTH, government hospitals, private hospitals, and primary health centres were classified as hospital-facility contacts. In contrast, home management by a doctor or nurse was classified as non-hospital. Pathway complexity was defined as the number of recorded steps, including the final arrival at UNIMEDTH, and was classified as one-, two-, three- or four-step pathways.

### Statistical analysis

Frequencies and percentages were used to describe sociodemographic characteristics, first points of contact, timing categories, and documented pathway sequences. The ten most frequently observed distinct sequences are reported, with all remaining sequences combined to preserve a readable summary of the heterogeneous routes. Because the time to presentation was right-skewed, the median time was compared between hospital-facility and non-hospital first-contact groups using the Mann-Whitney U test. The median times across the three grouped first-contact categories and the four pathway-complexity categories were compared using Kruskal-Wallis tests. Statistical significance was assessed at p < 0.05. Analyses were conducted using IBM SPSS Statistics version 29.

### Ethical considerations

Ethical approval was obtained from the University of Medical Sciences Teaching Hospital Research Ethics Committee before the extraction and analysis of the secondary records. Approval was granted on 17 July 2024 for the study titled “Determinant of time to presentation for stroke care in Ondo City.”

During data extraction from hospital records, the authors had access to information that could identify individual participants. However, all identifying information was removed after extraction, and the analytical dataset was anonymised before analysis. The anonymised dataset was stored securely and used only for the approved research purpose. Findings are reported in aggregate, with no information that could identify individual participants.

## Results

A total of 371 recorded stroke cases were included in the analysis. The mean age of patients was 59.86±14.17 years. Table 1 presents the sociodemographic composition of documented stroke cases extracted from hospital records at UNIMEDTH, Ondo State. The age distribution was concentrated in the middle and older adult age groups. Approximately half of the cases occurred among persons aged 45–64 years, while slightly more than one-third were recorded among those aged 65 years or older. By contrast, persons younger than 45 years accounted for only 14.3% of the recorded cases. The sex distribution was moderately male-dominated, with males accounting for 55.3% of the cases and females for the remainder.

**Table 1.**
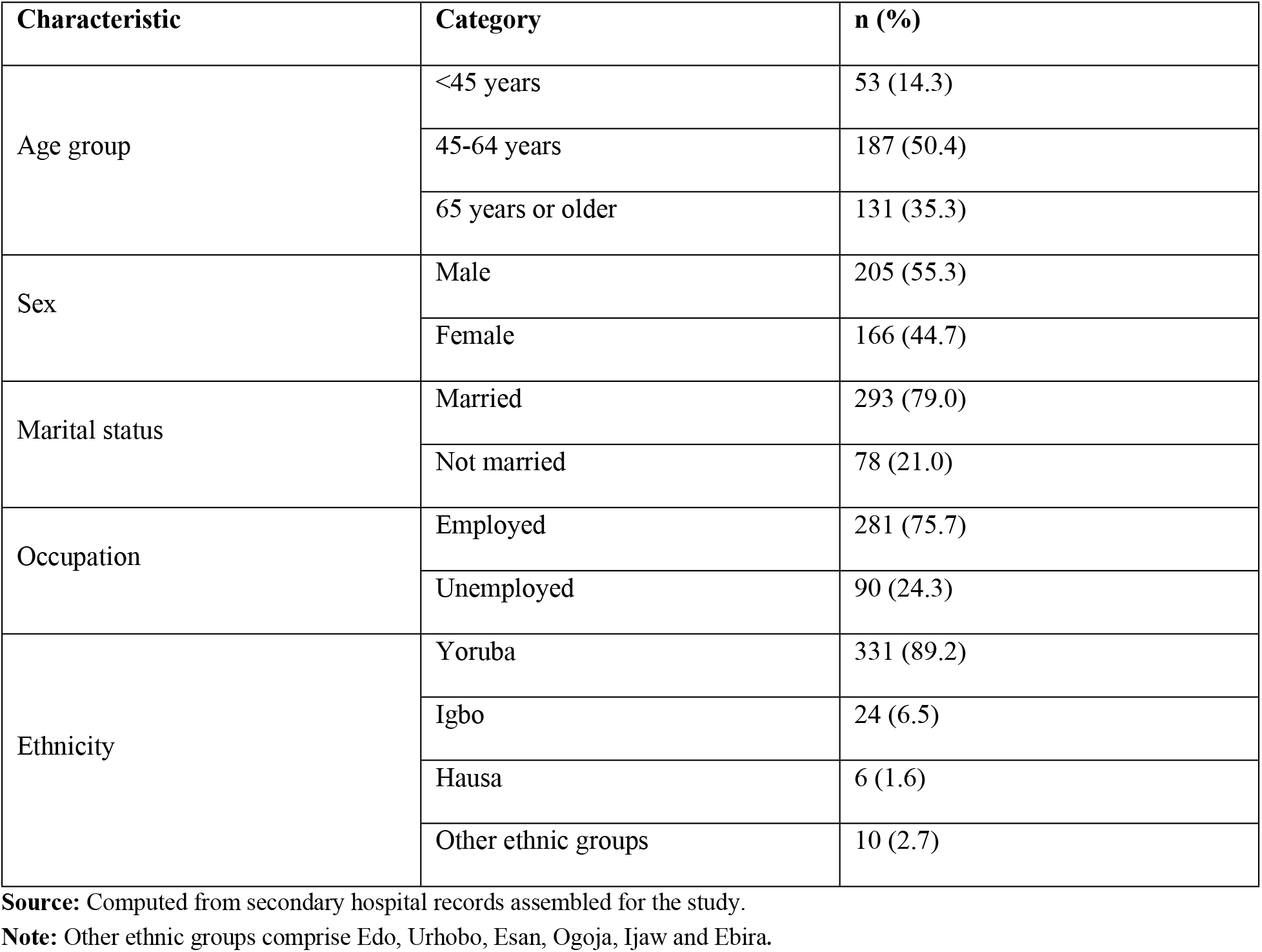
Sociodemographic characteristics of stroke cases identified from secondary hospital records at UNIMEDTH, Ondo State, Nigeria.

The marital-status distribution indicates that about 4 out of every 5 documented cases involved married persons, while approximately 1 in 5 involved unmarried persons. Regarding employment status, slightly more than three-quarters of the cases were recorded among employed persons, compared with 24.3% among those classified as unemployed. The ethnic composition was highly concentrated among Yoruba persons, who accounted for almost 9 out of every 10 documented cases. Igbo persons accounted for 6.5% of the cases, while Hausa persons and those from other recorded ethnic groups accounted for only a small proportion. Overall, the sociodemographic profile of the hospital-recorded stroke cases was characterised by a predominance of middle-aged and older adults, males, married persons, employed persons and Yoruba persons.

Table 2 presents the distribution of patients by their first point of contact after the recorded onset of stroke symptoms. Self/informal care was the most common first-contact category, accounting for 41.2% of documented pathways. Hospital/biomedical contact represented 31.0%, while slightly more than one-quarter of patients first sought traditional or faith-based care. Thus, initial contact occurred outside the hospital/biomedical category in more than two-thirds of recorded cases.

**Table 2.** First point of contact following recorded stroke symptom onset (n = 371).

| First-contact category | Specific first point of contact | n | % |
| --- | --- | --- | --- |
| **Hospital/biomedical** | **All hospital/biomedical contacts** | **115** | **31.0** |
|  | Managed at home by a doctor/nurse | 34 | 9.2 |
|  | UNIMEDTH | 30 | 8.1 |
|  | Private hospital | 23 | 6.2 |
|  | Government hospital | 20 | 5.4 |
|  | Primary health centre | 8 | 2.2 |
| **Self/informal** | **All self/informal contacts** | **153** | **41.2** |
|  | Chemist/patent medicine vendor | 83 | 22.4 |
|  | Self-medication | 70 | 18.9 |
| **Traditional/faith-based** | **All traditional/faith-based contacts** | **103** | **27.8** |
|  | Traditional home remedy | 71 | 19.1 |
|  | Traditional healer | 17 | 4.6 |
|  | Prayer house | 15 | 4.0 |

**Source:** Computed from secondary hospital records assembled for the study.
**Note:** The hospital/biomedical category includes management at home by a doctor or nurse as biomedical contact.

Within the hospital/biomedical category, management at home by a doctor or nurse was the most frequently recorded first contact, accounting for 9.2% of all patients. Direct presentation at UNIMEDTH accounted for 8.1%, followed by initial presentation at a private hospital and a government hospital. Primary health centres were the least frequently recorded hospital/biomedical first-contact point, accounting for 2.2% of the distribution. Among self/informal first-contact routes, chemist or patent medicine vendor use was more commonly recorded than self-medication, accounting for 22.4% and 18.9%, respectively. Within the traditional/faith-based category, traditional home remedies were the dominant initial contact, accounting for nearly one-fifth of all documented pathways. Traditional healer contact accounted for 4.6%, while prayer house contact represented 4.0%. Overall, the first-contact profile was characterised by a predominance of self- and informal routes, followed by hospital/biomedical and traditional/faith-based routes.

Table 3 presents the 10 most frequently observed distinct care-pathway sequences leading to UNIMEDTH. Nearly three-fifths of documented pathways were concentrated within these 10 routes, while 41.5% were distributed across the remaining 34 sequences. The most frequently recorded pathway was movement from a chemist or patent medicine vendor directly to UNIMEDTH, accounting for 10.2% of all documented sequences. This was followed by the pathway from traditional home remedies to a private hospital and, subsequently, to UNIMEDTH, which accounted for 8.4%. Direct presentation to UNIMEDTH ranked third, accounting for 8.1% of recorded pathways.

**Table 3.**
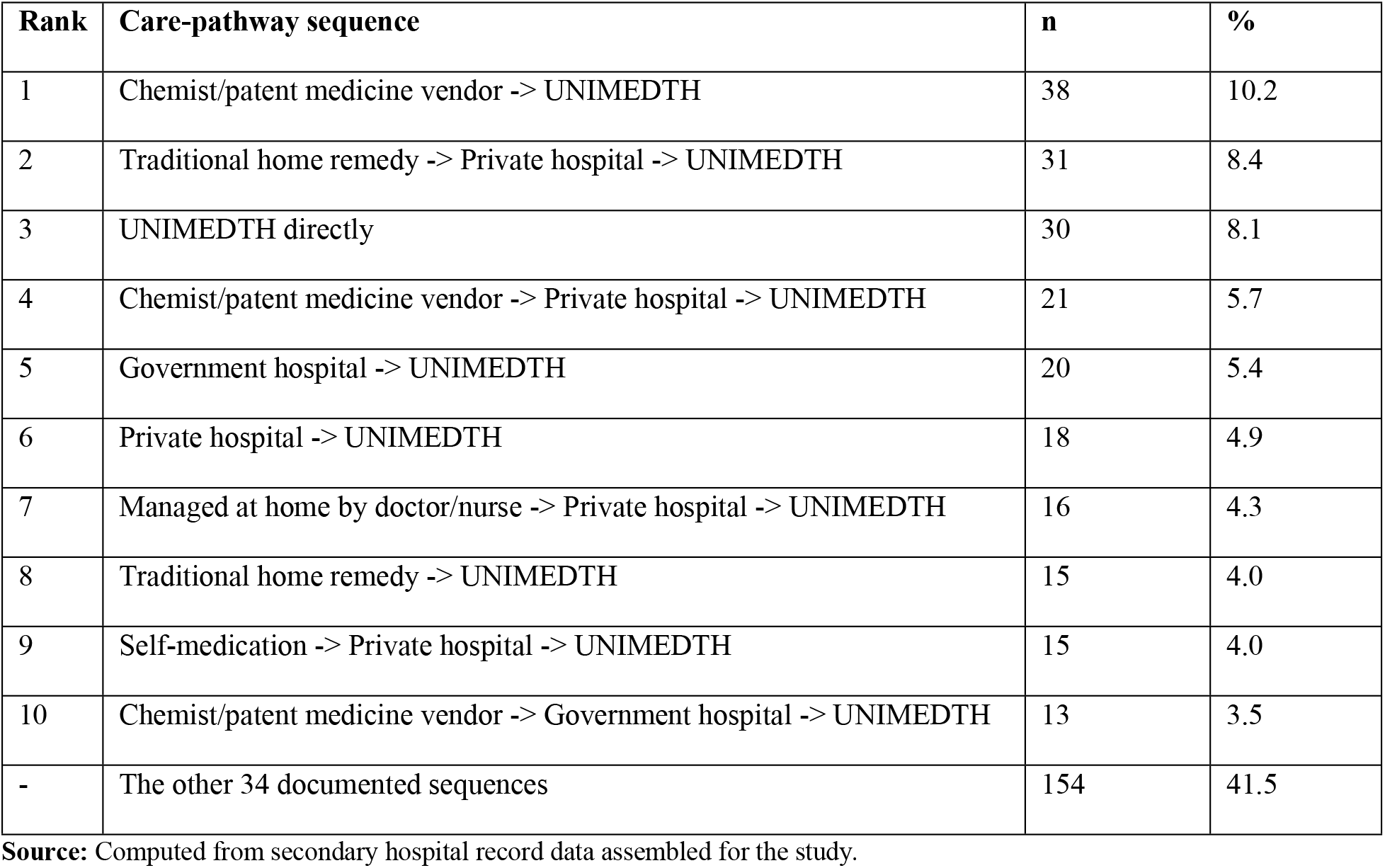
The 10 most frequently observed distinct care-pathway sequences leading to UNIMEDTH.

| Rank | Care-pathway sequence | n | % |
| --- | --- | --- | --- |
| 1 | Chemist/patent medicine vendor -> UNIMEDTH | 38 | 10.2 |
| 2 | Traditional home remedy -> Private hospital -> UNIMEDTH | 31 | 8.4 |
| 3 | UNIMEDTH directly | 30 | 8.1 |
| 4 | Chemist/patent medicine vendor -> Private hospital -> UNIMEDTH | 21 | 5.7 |
| 5 | Government hospital -> UNIMEDTH | 20 | 5.4 |
| 6 | Private hospital -> UNIMEDTH | 18 | 4.9 |
| 7 | Managed at home by doctor/nurse -> Private hospital -> UNIMEDTH | 16 | 4.3 |
| 8 | Traditional home remedy -> UNIMEDTH | 15 | 4.0 |
| 9 | Self-medication -> Private hospital -> UNIMEDTH | 15 | 4.0 |
| 10 | Chemist/patent medicine vendor -> Government hospital -> UNIMEDTH | 13 | 3.5 |
| - | The other 34 documented sequences | 154 | 41.5 |

**Source:** Computed from secondary hospital record data assembled for the study.

Among the remaining frequently observed sequences, the route from a chemist or patent medicine vendor via a private hospital to UNIMEDTH accounted for 5.7%. In comparison, presentation via a government hospital before arrival at UNIMEDTH accounted for 5.4%. A private hospital-to-UNIMEDTH pathway accounted for 4.9% of recorded sequences. Pathways involving initial management at home by a doctor or nurse, followed by presentation at a private hospital and subsequent arrival at UNIMEDTH, accounted for 4.3%. Traditional home remedies, followed directly by presentation at UNIMEDTH, and self-medication, followed by contact with a private hospital before arrival at UNIMEDTH, each accounted for 4.0% of documented routes. The tenth most frequently observed sequence involved contact with a chemist or patent medicine vendor, followed by a government hospital and then UNIMEDTH, accounting for 3.5%. Overall, the pathway distribution was spread across multiple routes, with the most common recorded sequences involving contact with a chemist or patent medicine vendor, a traditional home remedy, direct tertiary presentation, and intermediate hospital contact before arrival at UNIMEDTH.

Table 4 presents the distribution of time to tertiary hospital presentation across the recorded care-seeking pathway categories. Overall, the median time from recorded stroke symptom onset to presentation at the tertiary hospital was 24 hours, with an interquartile range of 9 to 72 hours. Under the specified timing classification, more than four in every five patients presented after four hours, while 14.6% presented within four hours of symptom onset.

**Table 4.** Care-seeking pathway categories and time to tertiary hospital presentation.

| Pathway measure | Category | n (%) | Median hours (IQR) | Test statistic | p-value |
| --- | --- | --- | --- | --- | --- |
| Overall presentation time | All stroke cases | 371 (100.0) | 24 (9-72) | - | - |
| Timing category | Early presentation ($\leq 4$ hours) | 54 (14.6) | - | - | - |
| | Late presentation ($> 4$ hours) | 317 (85.4) | - | - | - |
| Hospital-facility first contact | Hospital facility | 81 (21.8) | 3 (2-6) | U = 699.50 | $< 0.001$ |
|  | Non-hospital | 290 (78.2) | 48 (24-72) |  |  |
| Grouped first-contact category | Hospital/biomedical | 115 (31.0) | 6 (2.5-24) | H(2) = 99.92 | $< 0.001$ |
|  | Self/informal | 153 (41.2) | 24 (24-48) |  |  |
|  | Traditional/faith-based | 103 (27.8) | 48 (24-144) |  |  |
| Pathway complexity | One-step pathway | 30 (8.1) | 2 (1-3) | H(3) = 120.80 | $< 0.001$ |
|  | Two-step pathway | 129 (34.8) | 23 (6-48) |  |  |
|  | Three-step pathway | 184 (49.6) | 48 (24-72) |  |  |
|  | Four-step pathway | 28 (7.5) | 48 (24-150) |  |  |

**Source:** Computed from secondary hospital record data assembled for the study.
**Note:** IQR = interquartile range; U = Mann-Whitney U statistic; H = Kruskal-Wallis H statistic. Hospital-facility first contact identifies an initial encounter at a hospital facility.

Time to tertiary presentation varied substantially depending on whether the first recorded contact was with a hospital facility. Patients whose first contact was with a hospital facility had a median presentation time of 3 hours, compared with 48 hours for those whose first contact was outside a hospital facility. This difference was statistically significant (U = 699.50, p < 0.001). A graded pattern was also observed across the grouped first-contact categories. Patients whose first contact was within the hospital/biomedical category had the shortest median presentation time of 6 hours. The median time increased to 24 hours among those whose first contact was self- or informal care, and further to 48 hours among those whose first contact was traditional or faith-based care. The variation in presentation time across the first-contact categories was statistically significant, H(2) = 99.92, p < 0.001.

Presentation time also varied by pathway complexity. Patients with a one-step pathway had a median presentation time of 2 hours, whereas those with a two-step pathway had a median of 23 hours. Among patients with three- and four-step pathways, the median rose to 48 hours. The four-step pathway also had the widest interquartile range, spanning 24 to 150 hours. The difference in presentation time across levels of pathway complexity was statistically significant, H(3) = 120.80, p < 0.001. Overall, the table shows progressively longer median presentation times across non-hospital first contacts, traditional or faith-based routes, and more complex care-seeking pathways.

## Discussion

This study examined documented routes to tertiary stroke care rather than treating arrival time solely as an individual delay outcome. Three findings stand out. First, direct presentation to UNIMEDTH was uncommon: fewer than one in ten patients reached the tertiary hospital as their first recorded care contact. Second, most patients initially entered care through routes outside a hospital facility, especially chemists, patent medicine vendors, self-medication, and traditional home remedies. Third, these pathway patterns were strongly associated with time to tertiary arrival: patients who first contacted a hospital facility arrived substantially earlier than those who entered through non-hospital routes. In contrast, multi-step pathways were associated with progressively longer presentation times.

The median presentation time of 24 hours and the high proportion presenting after four hours are consistent with the broader African evidence base. Earlier reviews reported long onset-to-admission intervals and major deficiencies in recognition, transport and organised stroke services across the continent [14-20]. More recent synthesis also demonstrates that prehospital delay remains widespread in Africa [7]. The present study extends this literature by showing the route embedded within delay: most patients recorded at UNIMEDTH had already engaged with some form of care, but the care first accessed frequently did not place them on a rapid tertiary stroke pathway.

The finding that chemists or patent medicine vendors were the most common specific first-contact point is particularly consequential. These vendors occupy an established position in Nigerian care-seeking because they are geographically accessible, familiar and often available with fewer barriers than formal facilities [13]. Their prominence in the present pathways should therefore not be read simply as inappropriate patient behaviour. Rather, it identifies a segment of the health system that may be critical for rapid symptom recognition and referral. In the context of stroke, a provider who can rapidly recognise sudden facial asymmetry, arm weakness or speech disturbance and immediately refer rather than dispense medication or observe treatment response may shorten the time lost before definitive assessment.

The presence of traditional home remedies, traditional healers and prayer houses in the documented routes is also consistent with the social plurality of care in Nigeria. This is in tandem with the findings of a qualitative study that found that stroke survivors may move across care settings along complex therapeutic itineraries before reaching rehabilitation [7]. Kleinman’s framework helps explain why such routes may remain meaningful to patients and households: stroke symptoms may be interpreted through biomedical, spiritual and social meanings simultaneously [26]. Accordingly, interventions that merely denounce non-biomedical care are unlikely to be sufficient. A more pragmatic response is to engage traditional and faith-based actors as partners in urgent referral for suspected stroke, while respecting their continuing social role in support and meaning-making.

The significant temporal gradient across pathway complexity is equally important. Patients presenting directly to UNIMEDTH had a median arrival time of two hours; those moving through two or more steps generally presented much later. These additional steps may represent treatment trials, reassessment after symptoms persist, household deliberations, transport organisation or referrals between providers. Although the record data do not disaggregate the duration spent at each step, the pattern suggests that the transitions themselves offer opportunities to reduce delay. Similar concerns arise from evidence in Uganda, where many patients reaching national referral hospitals had initially attended lower-level facilities and experienced prehospital delay [16].

The findings have direct implications for service delivery. Public messaging remains necessary because symptom recognition is the first step in urgent response; however, community awareness alone cannot address delays, as most patients initially contact providers outside tertiary hospitals. The World Stroke Organisation statement on LMIC prehospital care calls for coordinated action encompassing awareness, emergency transport, and the engagement of primary, traditional, and faith-based providers [9]. In Ondo State, locally feasible actions may include brief stroke recognition and immediate referral protocols for patent medicine vendors and community-facing providers, referral linkages between private or lower-level facilities and UNIMEDTH, defined emergency communication channels, and routine documentation of onset time and referral transitions.

The theoretical interpretation is also useful. Andersen’s model suggests that initial provider choice may be shaped by accessibility, affordability, available transport and perceived urgency, even where the condition eventually requires tertiary care [23,24]. The pathway framework, therefore, moves the analysis away from an assumption that late presentation necessarily reflects unwillingness to seek treatment. Rather, delayed arrival may occur because people seek the form of care that is available, trusted or understood locally, but encounter weak linkages to definitive stroke services. This is precisely the interface at which system redesign can intervene.

This study has strengths and limitations. Drawing on the stroke registry, radiology records, referral notes and ambulance records made it possible to reconstruct documented pathways rather than relying only on a single contact field. Nevertheless, the retrospective design depends on the completeness and accuracy of recorded symptom-onset times and provider contacts. The analysis includes only patients who ultimately reached UNIMEDTH and therefore cannot characterise individuals who remained in community care, died before referral or attended other hospitals. The study also did not estimate independent associations after adjustment for stroke severity, distance, socioeconomic resources, transport mode or other potential confounding factors. The non-parametric comparisons should therefore be interpreted as evidence of temporal differences across recorded pathways, not as proof that particular care choices caused delay.

## Conclusion

Care-seeking pathways to tertiary stroke care in Ondo State were heterogeneous and seldom direct. Most recorded patients first used a non-hospital route or passed through multiple contacts before arriving at UNIMEDTH, and these pathways were associated with markedly longer time to presentation. Delayed tertiary presentation should therefore be understood partly as a pathway and referral system problem, rather than solely as delayed help-seeking by patients or families. A credible stroke-delay reduction strategy in this setting must combine community awareness with rapid integration of referrals across hospitals, chemists and patent medicine vendors, traditional and faith-based providers, and emergency transport services.

## Data Availability

The data used for this study were extracted from clinical and referral records and may contain potentially identifying patient information. A final data availability statement, including the applicable institutional access procedure and contact point, should be confirmed with UNIMEDTH and its Research Ethics Committee before submission.

